# SAHVAI-3D and 4D: A New, Automated Subarachnoid Hemorrhage Volumetric Artificial Intelligence (SAHVAI) Measurement Approach Using Non-Contrast Head CT Scans

**DOI:** 10.1101/2024.08.29.24312799

**Authors:** Melina Wirtz, Saif Salman, Yujia Wei, Vishal Patel, Rohan Sharma, Vikash Gupta, Qiangqiang Gu, Benoit Dherin, Sanjana Reddy, Rabih Tawk, Bradley J Erickson, William David Freeman

## Abstract

**Objectives:** To automate subarachnoid hemorrhage volume (SAHV) calculation (***SAHVAI-SAHV A****rtificial* ***I****ntelligence)* and create 3D volumetric images (SAHVAI-3D) using non-contrast head CT (NCCT) imaging data in aneurysmal subarachnoid hemorrhage (SAH) patients. We also defined SAHVAI-4D, representing SAHV over time. The aim was to compare automated SAHVAI volumes to manual SAHV methods and computation times, explore these imaging biomarkers’ potential in identifying at-risk brain regions for delayed cerebral ischemia (DCI), and explore potential insights in future neurotherapeutic interventions for SAH patient recovery.

**Methods:** A training set of 10 consecutive aneurysmal SAH cases was used to manually compute SAHV, SAHVAI-3D, and SAHVAI-4D, involving 92 non-contrast CT scans (182 slices each). The SAHVAI deep learning (DL) algorithm generated automated SAHV values in cubic centimeters (cc). For both SAHVAI and manual evaluations, a 3D SAH brain map was created for each patient. Blood volumetric outputs were analyzed and compared to neurological outcomes at discharge, including DCI events, symptomatic vasospasm (sVSP), and areas with the thickest SAH blood concentration.

**Results:** SAHVAI quantified SAH blood volume (SAHV) in average of 6.7 seconds per scan, significantly faster than the manual method, which took over 60 minutes per scan (Fisher’s exact test, P value <0.001). SAHVAI demonstrated an accuracy of 99.8%, a Dice score of 0.701, a false positive rate of 0.0005, and a negative predictive value of 0.999. The mean absolute error between SAHVAI and manual methods was 5.67 ml. The SAHVAI-3D brain map and total SAHV at admission were strongly associated with neurological outcomes, inversely with Glasgow coma scale (R^2^=0.23, p=0.017) and directly with length of hospital stay (R^2^=0.175, p=0.004), especially in regions with dense blood concentration.

**Conclusion:** SAHVAI-3D and SAHVAI-4D brain mapping techniques represent innovative imaging biomarkers for SAH. These advancements enable rapid evaluation and targeted interventions, potentially improving patient care in SAH management.

## Introduction

Aneurysmal subarachnoid hemorrhage (aSAH) is a disproportionately deadly stroke subtype, accounting for up to 3% of all strokes^1^ but with a 40% one-month mortality rate. The global incidence of subarachnoid hemorrhage (SAH) is approximately 6.1 per 100,000 person-years^2^. Despite a 1.7% annual increase in aSAH incidence from 1955 to 2014^2^, the disease remains highly fatal, with median mortality rates reported at 32% in the US, 44% in Europe, and 27% in Japan^3^. However, between 1980 and 2020, the global case fatality rate for SAH declined by 1.5% annually due to advancements in stroke care systems and neurologic intensive care units^4^. These units utilize advanced neurodiagnostics such as lumbar puncture, CT angiography, MRI, MR angiography, and digital subtraction cerebral angiography to diagnose, monitor, and treat acute SAH patients^5^. The incidence of SAH increases with age^6^, especially in women^7^, who are 1.24 times more likely to be affected than men. With an aging population, the healthcare burden of SAH continues to grow^6^. Prompt detection and management of SAH in emergency departments are critical for patient outcomes^8,9^. Diagnosing aSAH can be challenging because its symptoms, such as headache, can mimic those of ischemic stroke^9,10^. Non-contrast CT (NCCT) scans remain the gold standard for confirming suspected SAH^10^. Management of SAH is complicated by common issues such as rebleeding, increased intracranial pressure, seizures, fever, hypothermia, cardiopulmonary complications, hyponatremia, deep venous thrombosis, hydrocephalus, vasospasm (VSP) and delayed cerebral ischemia (DCI)^11,12^. Among survivors, about 30% have moderate to severe disability, and approximately 66% of those who undergo successful neurosurgical clipping of the aneurysm do not regain their pre-hemorrhage quality of life^10,13^. DCI, which affects 30% to 40% of SAH patients, is associated with poor outcomes and remains difficult to predict and treat^14,15^. Research by Sharma et al. has shown correlations between patient age, Glasgow Coma Scale (GCS) scores, and SAH blood volume (SAHV) with discharge outcomes, prediction of DCI during hospitalization, and in-hospital mortality^16^. This study hypothesizes a direct *dose-response* relationship between initial SAHV, its clearance over time, and clinical outcomes. To improve measurement and visualization of SAHV, we developed a deep learning (DL) method ^17^, SAHVAI, which automatically segments SAHV from NCCT head data. This method aims to provide rapid and precise measurements of SAHV in both 2D axial CT slices and a 3D brain map, improving visualization of blood volume and location. The DL approach uses ubiquitous NCCT scan data for diagnosing SAH blood and monitoring postoperative conditions such as external ventricular drain (EVD) placements and/or other neurosurgical procedures for monitoring rebleeding and basic brain structure information in terms of the Hounsfield units and windows (brain, bone, soft tissue, etc.). SAHVAI-3D, aims to provide a precise diagnostic parameter at admission, potentially surpassing the modified Fisher scale, which is semi-quantitative.

These approaches could serve as novel neuroimaging biomarkers for SAH patients, aiding drug discovery and neurosurgical interventions. To validate SAHVAI, we developed a manual method (MM) where a human investigator segmented and analyzed the NCCT scan images, considering this the gold standard for comparison. Volume measurements from the AI method and MM were linked and compared with clinical data to explore the dose-response relationship between aSAH blood volume and neurological outcomes within the first 30 days post-hemorrhage.

## Methods

### Demographic data

Our cohort included 10 patients with aSAH treated from March 2019 to February 2022. The cases were chosen from an existing Mayo Clinic secure RedCap database and further clinical information of interest was collected from Epic Electronic health Records (EHRs) to cross-match with the RedCap data. NCCT scans of the head performed for standard of care indications were utilized during the patient’s hospital stay for analysis. When more than one CT scan was performed in a single day, the earliest NCCT performed that day was always utilized. Among this cohort of 10 patients, there were 7 females and 3 males; the average age was 55 yearsat SAH. Admission weight was reported as an average of 92 kg (minimum 55 kg, maximum 120 kg).

In all ten cases, the following data have been collected from our cohort and were analyzed about SAHV over time: age at SAH, gender, ethnicity, race, type of SAH, admission weight, modified Fisher score, GCS on admission, Hunt and Hess on admission, World Federation of Neurological Surgeons scale, National Institutes of Health Stroke Scale (NIHSS) at admission, Physiologic Derangement Scale at admission, external ventricular drainage placed, aneurysm surgery, angiographic vasospasm (VSP) (Transcranial doppler ultrasound (TCD), computed tomography angiography (CTA) or both), day of VSP, severity of VSP, location of VSP if obtainable, management of VSP, symptomatic clinical VSP, new cerebral infarction on imaging, description of infarct, DCI, day of DCI, re-rupture/rebleeding, pre-listed modified Rankin Scale (mRS), mRS at discharge, mRS at 30 days, mRS at 90 days, SAH associated disease, length of stay in hospital (LOS), length of stay in intensive care unit, hypertension, diabetes mellitus, heart disease, pure motor hemiparesis stroke, smoker, alcoholic, and family history of aneurysm.

### SAHVAI-Subarachnoid Hemorrhage Volumetric Artificial Intelligence Method

An DL-driven approach to manual SAH segmentation was developed to increase throughput and repeatability. After IRB approval, we utilized our SAHVAI model trained with U-Net architecture on 150 manually SAH-labeled NCCT ground truth labeled images to create SAHVAI-3D and 4D models on an NVIDIA V100 32G GPU^33^. The resulting model was then applied in inference mode to the same cohort of the 10 patients and their 92 NCCT scans. Using both manual method data sets (MM-SAHV) and SAHVAI volumes, we then compared them for measurement differences in cc volumes. We also compared 2D and 3D SAHV brain maps (**Figure 1**) to visualize SAH blood in 3D (similar to Maximum Intensity Projection (MIPS) maps in radiology). Compared to the MM which measured eight spaces, SAHVAI visualize five major cisternal SAH anatomical blood spaces.

**Figure 1.**
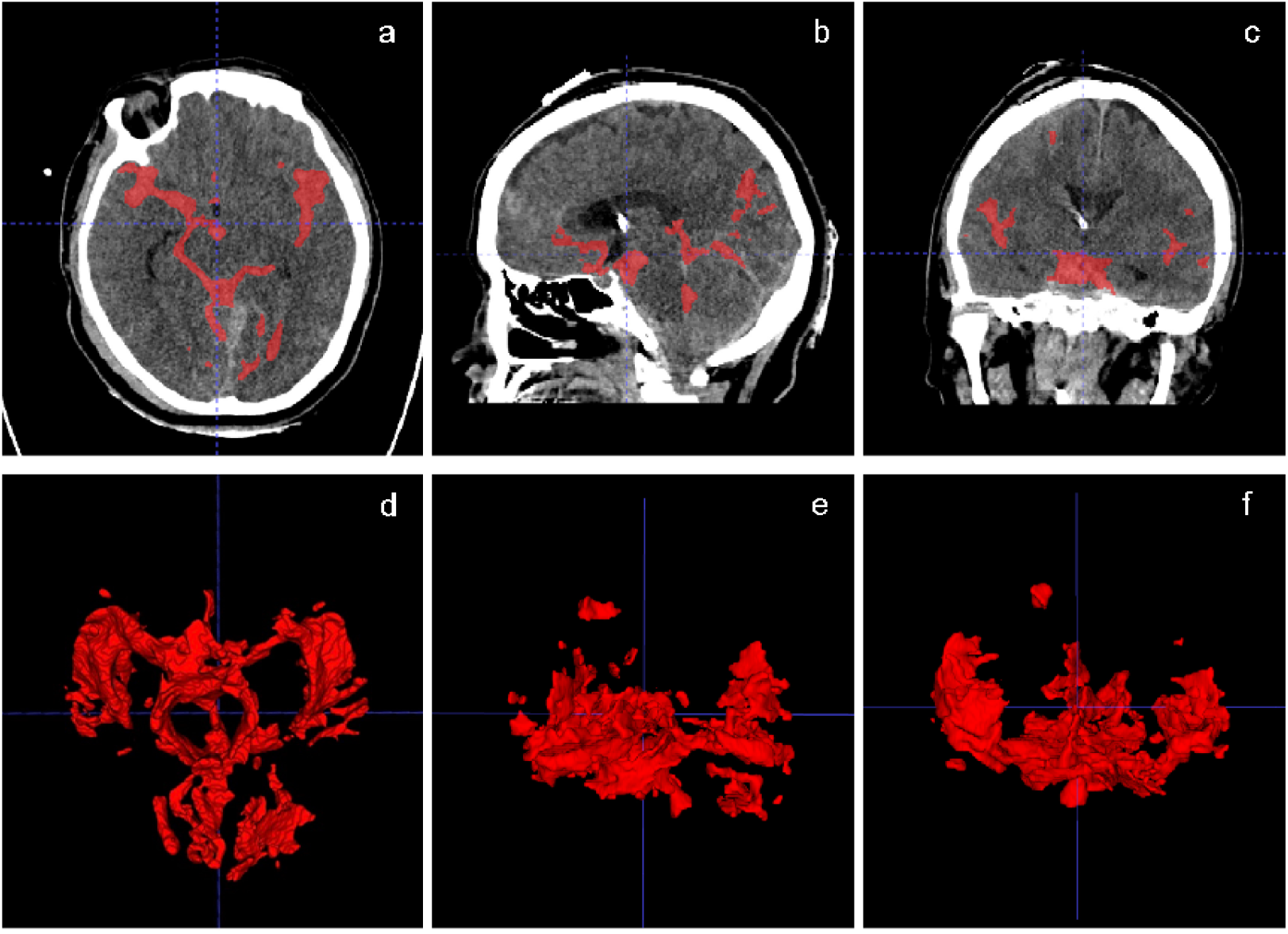
End-result SAHVAI Artificial Intelligence of SAHV in 2-dimensional (a, b, c) and 3-dimensional (d, e, f) SAHV-3D Brain Map formats. This SAHVAI-3D Brain Map is from the first case on day 0 (incident day of SAH). Segmentation of five cisternal spaces in red equals blood. Planes: axial (a); sagittal (b); coronal (c); View from axial (d); sagittal (e); coronal (f).

### Manual Method of SAHV Measurement

Based on the patient cohort, we developed a new manual method (MM) to measure the SAHV in each CT scan, which was considered the gold standard reference value (i.e., ground truth) compared with the results of the SAHVAI method. We used ITK-SNAP (4.0) software to apply the new MM and started with an intensity-based segmentation, selecting a voxel with Hounsfield Units (HU) between 60 and 120. Then, using a window center of 60 HU and width of 120 HU, the intensity-based segmentation was manually refined with attention to eight defined neuroanatomical spaces (five cisternal spaces: suprasellar cistern, perimesencephalic cistern, prepontine cistern, sylvian cistern, interhemispheric cistern), and three additional neuroanatomical spaces relevant to SAH disease: intraparenchymal hemorrhage (IPH), intraventricular hemorrhage (IVH), and finally brain gyral/sulcal spaces on each slice. An example of the final result is shown in **Figure 2** as a 2D (upper images) and 3D (lower images) called the SAHV-3D, also called “SAHV-3D Brain Map”. These 3D images can be rotated like any other reconstruction image used in neurosurgery or neurointerventional to visualize patterns in 3 dimensions, which is hard to visualize in standard 2D axial planes.

**Figure 2.**
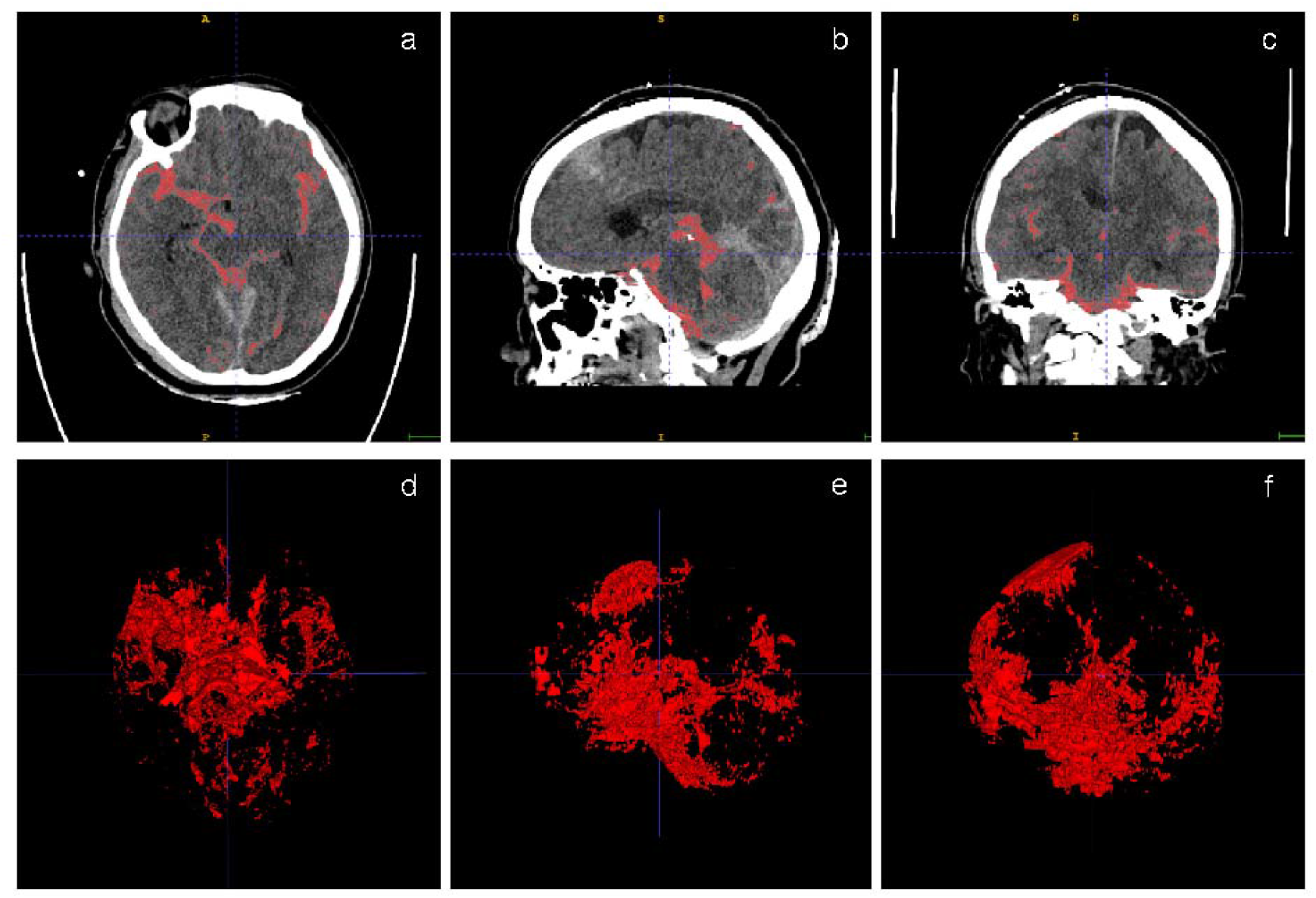
End-result of SAHV Manual Method in 2-dimensional (a, b, c) and SAHV 3D Brain Map (d, e, f) subarachnoid hemorrhage volume. This SAHV-3D Brain Map is from incident day of SAH. Segmentation of eight spaces (five cisternal spaces, Intraparenchymal Hemorrhage (IPH), Intraventricular Hemorrhage (IVH), and gyral/sulcal spaces) in red equals blood. Planes: axial (a); sagittal (b); coronal (c); View from axial (d); sagittal (e); coronal (f).

The final segmentation also yielded the quantified SAH volume (qvSAH) or SAHV via a straightforward multiplication of the total SAH voxel count and single voxel volume.

### Statistics and Visualization

The Manual and the AI Method brain maps were analyzed and graphically compared to neurological outcomes at 30 days by mRS and visual areas of thickest blood concentration concerning DCI and symptomatic vasospasm (sVSP) events.

**Figure 3 and Table 1** summarize all descriptive statistics and the regression analysis of SAHV and neurological outcomes.

**Table 1.**
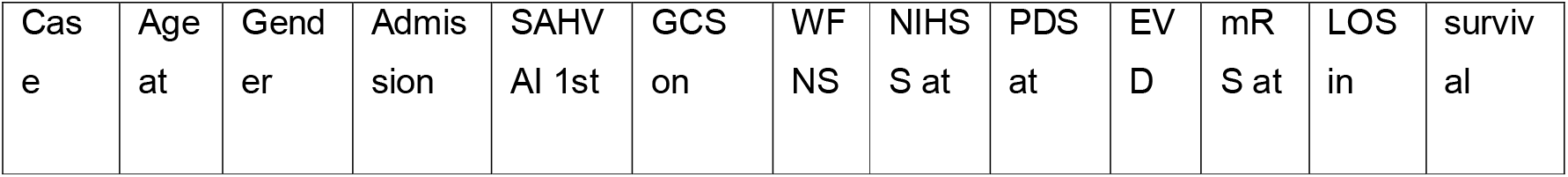
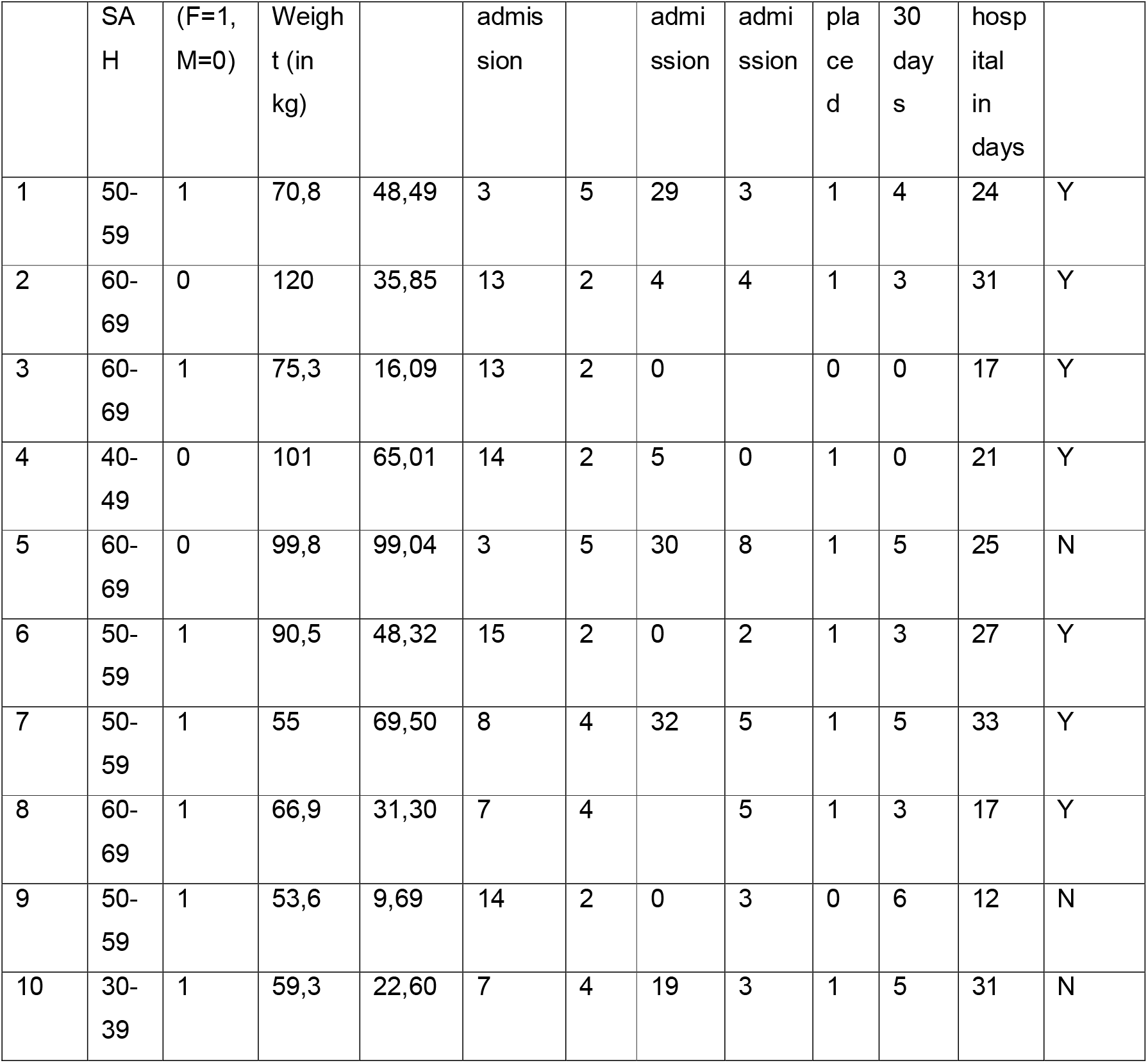
Quantitative SAH Volume (qvSAH) measured by the AI method of the first CT scan compared to neurological outcome (Neurological Examination Scores and LOS in hospital). Abbreviations: NIHSS = NIH Stroke Scale/ Score; PDS = SAH-Physiologic Derangement Scale; EVD = external ventricular drainage.

**Figure 3.**
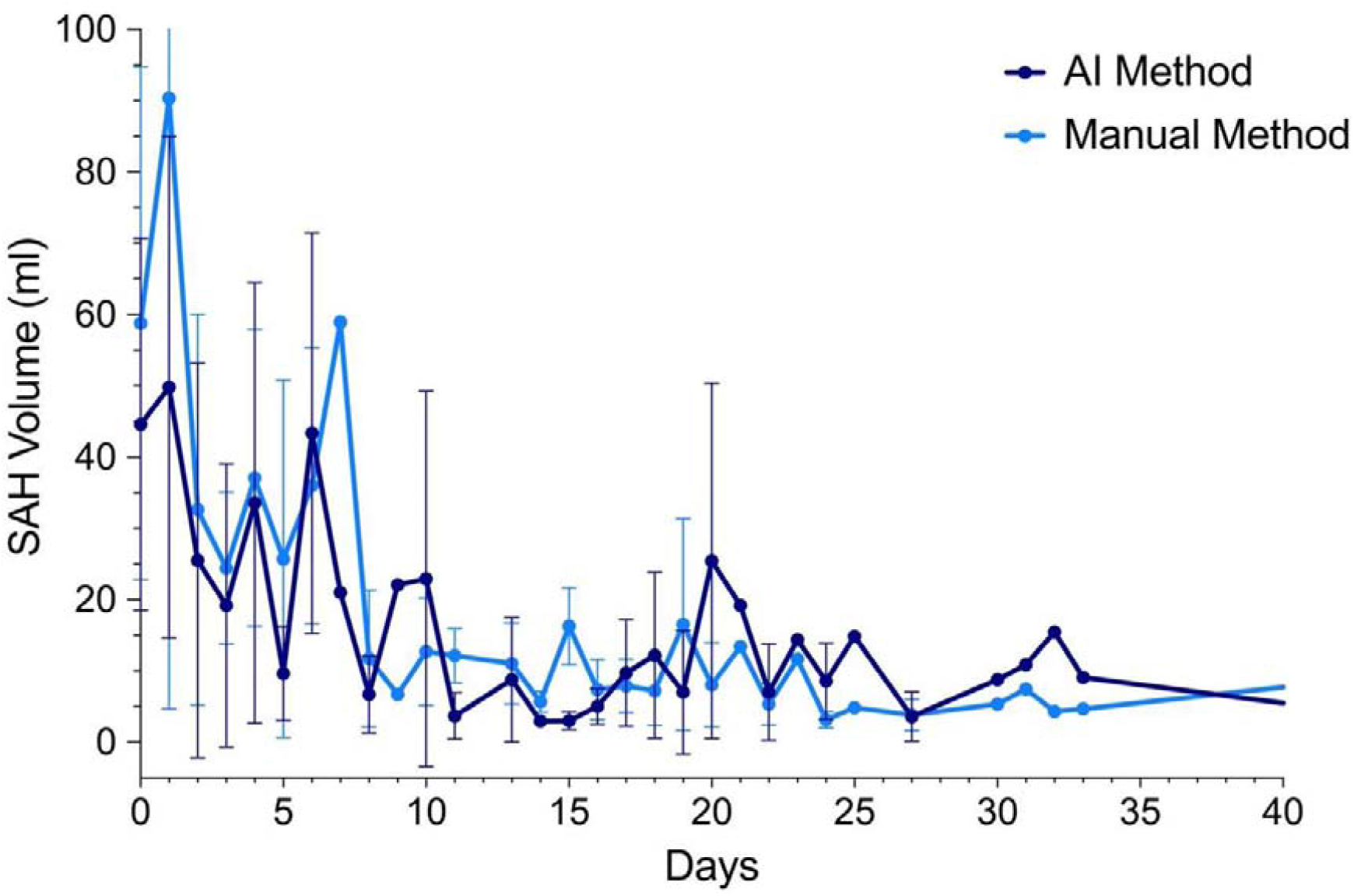
Plot of the Mean and Standard Deviation of AI (dark blue) and Manual (light blue) Methods measured the quantitative SAH volume (SAHV) over time of the total cohort (n=10) day by day.

## Results

For 3D visualization, we analyzed 92 CT scans to measure SAHV blood. A minimum of 5 CT scans/cases were analyzed, and a maximum of 13 CT scans/cases were observed in the 10 patient 3D, 4D cohort. On average, 9.2 CT scans were studied for each patient. We only included SAH cases with a subsequent NCCT after the initial SAH ictus or admission scan. The length of stay (LOS) of all ten SAH patients was on average 23.8 days, whereas the average day of the last CT scan included in this method was on day 29.44 after the first CT scan and, therefore, post-ictus. The shortest duration between the first and last CT scans evaluated in this project was 11 days; however, this patient died on day 16. Otherwise, the most extended duration between day zero (the day of the incident) and the last CT scan included was 69 days. Of all 92 CT scans, the average axial CT slices were 181.61 for each CT scan (minimum 64 slices, maximum 272 slices). So, in total, 16708 CT slices were examined regarding SAH blood volume using these methods.

### SAHVAI-4D: Mean and Standard Deviation of SAHV over Time (AI and Manual Methods)

We analyzed the SAH Volume over time of all 10 patients and their NCCT. A plot of the mean and standard deviation of Manual SAHV and SAHVAI methods (**Figure 3**) shows the natural decline of SAHV over time with both methods.

The decrease of SAHV over time seems likely consistent with clinical observations of SAH blood decline on NCCT. However, current methods typically use the Fisher scale, which is semi-quantitative, or other similar semi-quantitative methods like the Hijdra scale. Therefore, both SAHV methods demonstrate important data of maximal SAH blood happening within the first few days up until about 10 days post-ictus before flattening out. Around post-SAH day 15, the MM showed more SAHV than the SAHVAI automated method. But when we measured SAHV over time, we saw a switch around day 20 onwards) where the AI appears to slightly overfit manual evaluation blood measurement. The mean SAHV of day 0 or the day of the SAH ictus aneurysm rupture was measured as 44.59 ml by SAHVAI, whereas 58.78 ml by MM. The maximum SAHV measured for day 0 by SAHVAI was 99.01ml, and by SAHV MM, it was 141.5ml for both of the same cases (case 5). Whereas the minimum SAHV on day 0 measured by SAHVAI was 9.69ml (case 9) and MM was 12.8ml (case 10)). In **Figure 3**, the standard deviation (SD) of day 1 of the MM exceeds the selected scale unit. The SD of day 1 of the MM was calculated with 85.61, as the lowest measured SAHV was 10 ml (case 2), and the highest SAHV was 276.7 ml (case 9) on day 1. In general, one has to notice that the SD is high, as all ten cases have different severity of SAH bleeding. Taking the average standard deviation of each calculated mean SAHV of all CT scans, including the ten patients, AI has a mean SD of 9.09, and Manual has a mean SD of 9.32. On average, the last CT scans were 29.44 days after the aSAH rupture. However, two NCCT scans (case 2 with 69 days and case 3 with 47 days post-ictus) were included but are not displayed in **Figure 3**. For the sake of completeness, case 2 had, on day 69 after the incident, a SAHV of AI 0.7ml and Manual 1.3ml, and case 3 had, on day 47 after the SAH, a SAHV of AI 1.89ml and Manual 10.75ml.

### Absolute and Relative Differences between AI and Manual Methods

The mean absolute difference from day to day between the Manual and AI Methods of all ten cases was calculated as 5.76ml. This small volume of about 5ml is about the volume of one teaspoon, and we consider this to be notable but of uncertain clinical significance.^20^ The day-to-day absolute difference′s variance of the MM was more extensive than that of the AI method. The MM′s absolute difference varied in a range of 57.39ml (from 0.33ml to 57.72ml), whereas the AI method′s variance was 33.69ml (between 0.02ml to 33.71ml). Therefore, the overall variance in MM SAHV appears higher than the automated SAHVAI method. The highest absolute difference, and therefore the most clearance of SAHV, measured with the AI Method, was 33.71ml from day 5 to day 6 after the ictus. Meanwhile, the MM′s highest absolute difference was between day 1 and day 2, with 57.72ml. The day-by-day average relative difference between both methods was 25.7% (0.26).

### SAHVAI Model Performance

Regarding the SAHVAI Model Performance, SAHVAI, as an NCCT scan AI approach, rapidly quantifies the volume of aneurysmal SAH blood in approximately 6.7 seconds for 30 NCCT slices and approximately 42 seconds for 188 NCCT slices. In contrast, labeling the SAH blood volume with the manual method took 60 to 93 minutes for an average of 188 NCCT slices. The SAHVAI model currently needs approximately 15 GB of GPU memory and can run very smoothly on one NVIDIA V100 GPU. Furthermore, the SAHVAI achieved an accuracy of 99.8%, dice score of 0.701, FPR=0.0005, and NPV=0.999.

### Quantitative SAHV Compared to Neurological Outcomes and Associations

The SAHVAI-3D brain maps and quantified SAH volume (SAHV) at admission of all cohort patients also appeared highly associated with functional neurological outcomes (**Table 1**), especially regarding GCS and LOS in the hospital.

There might be a correlation between quantitative SAH blood volume (SAHVAI), GCS (linear regression R2=0.23, p=0.017; inverse association), and the LOS in the hospital (linear regression R2=0.175, p=0.004). Interestingly, we observed that the higher the SAHV in cc/ml, the lower the GCS score and the longer the patient′s LOS within the hospital, which is a novel discovery. We also saw a fascinating reciprocal observation that the lower the SAHV, the higher the GCS score and the shorter the patient’s LOS in the hospital. Because of the limited 3D data analysis of only 10 cases with 16708 CT slices with 10 mRS variables, we could not find a significant correlation between SAHV and the mRS and likely grossly underpowered. We plan to validate SAHV on mRS outcomes in our original 150 SAH training set.

### Comparing Results of 2-dimensional and 3-dimensional SAHV Brain Map

To illustrate the difference between the two methods (AI and MM), **Figure 4**. shows the 2-dimensional SAHV Brain Map of cases 1, 5, and 8.

**Figure 4.**
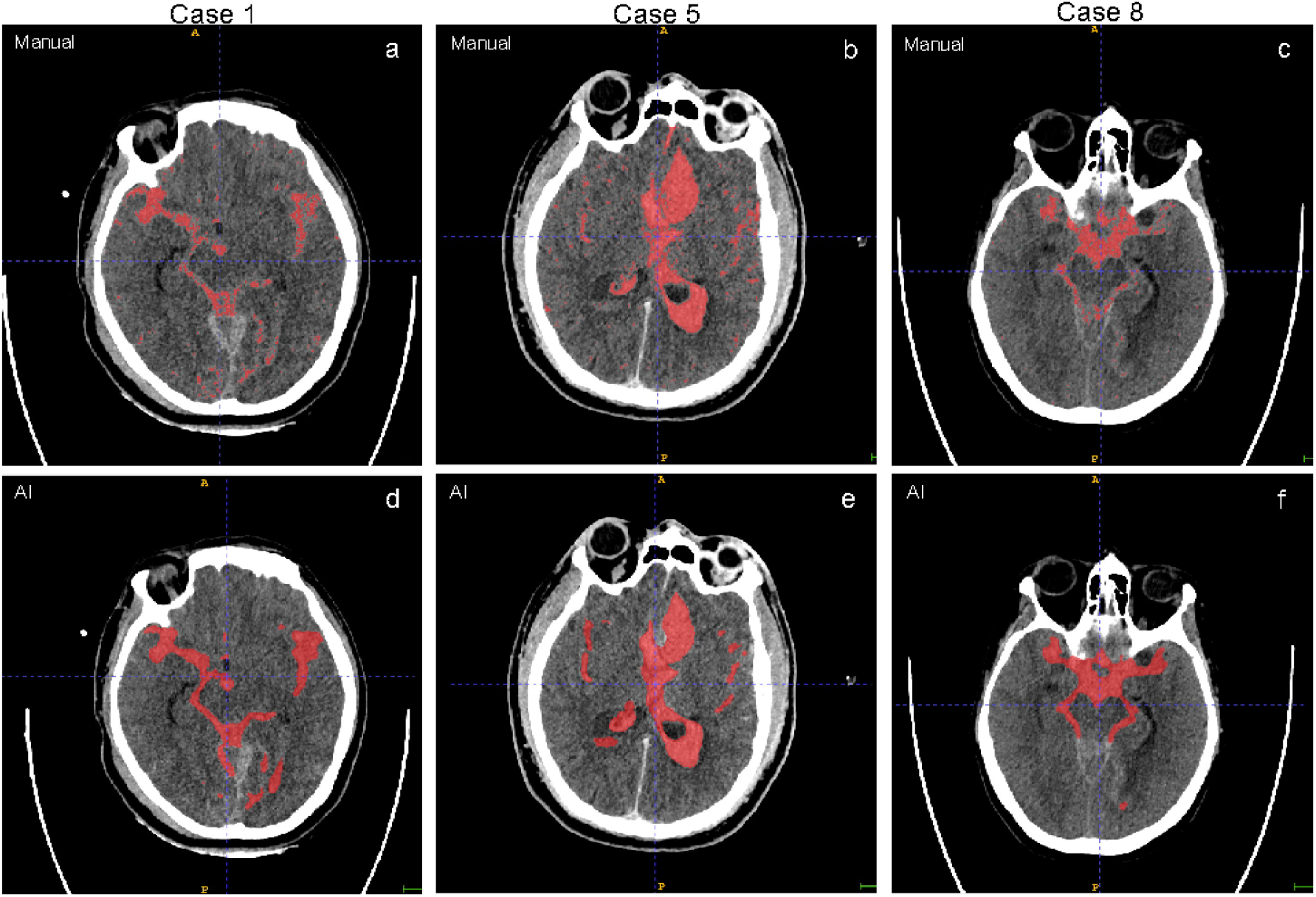
2-dimensional SAHV Brain Map of three cases (Case 1, Case 5, and Case 8) of Manual (a, b, c) versus SAHVAI (d, e, f) methods. All planes are axial. The MM segmented eight spaces are colored red as SAH blood, whereas the SAHVAI method labeled five spaces in red, which equals the SAH blood. Each NCCT scan is labeled with an overall opacity of 50%.

Paying attention to the SAHV-3D brain map of both methods (**Figures 1 and 2**) of the same patient (case 1) from day 0, with whom the patient was diagnosed with an aSAH, a visual area of the thickest blood concentration was seen in the right convexity of the brain. The VSP of this patient, which was diagnosed via TCD, CTA, and digital subtraction angiography (DSA), was symptomatic on the right middle cerebral artery (= location of visual area of thickest blood concentration in SAHVAI-3D). This tendency of the visible area of densest blood concentration, seen in both SAHV-3D brain maps (Manual and AI), is not only correlated in our first case with the later location of sVSP but also in additional cases of our cohort (**Table 2**).

**Table 2.** Severity and location of vasospasm (if obtainable) compared with the visual area of thickest blood concentration in SAHV-3D Brain Map (Manual and AI Method) of all ten cases. Abbreviations: VSP = Vasospasm; MCA = Middle Cerebral Artery; ACA = Anterior Cerebral Artery; VA = Vertebral Artery

| Case | Severity of Vasospasm | Location of VSP if obtainable | Visual area of thickest blood concentration on in SAHV-3D Brain Map |
| --- | --- | --- | --- |
| 1 | severe | right MCA | right convexity of the brain |
| 2 | N/A | N/A | diffuse |
| 3 | mild | right MCA | right convexity of the brain |
| 4 | severe | right MCA | right convexity of the brain |
| 5 | severe | diffuse | posterior and right convexity of the brain |
| 6 | severe | bilateral ACA | left convexity of the brain (upper part of the brain) |
| 7 | moderate - severe | diffuse | left convexity of the brain |
| 8 | mild | right VA | posterior location of the brain (brainstem) |
| 9 | severe | bilateral MCA, ACA, diffuse | diffuse, especially right convexity of the brain |
| 10 | mild | diffuse | diffuse |

## Discussion

Prior studies have shown that as the modified Fisher scale increases, which is a semiquantitative scale of SAH blood volume, it influences future risk symptomatic vasospasm.^16,21–23^ However, the modified Fisher scale faces challenges in predicting other important patient outcomes, such as a shunt or EVD dependency, and identifying specific brain tissue areas at risk. To our knowledge, this study is the first to report a novel deep learning (DL) and comparative manual approach that precisely and quickly measures SAH blood volume (SAHV) over time.

Our study demonstrates that as SAHV increases, it correlates inversely with the Glasgow Coma Scale (GCS) at admission and proportionately with the length of stay (LOS). Preliminary 3D topographic map data suggest that higher SAHV is associated with future severe symptomatic vasospasm risk. We compared the SAHVAI approach with a more tedious and time-intensive Manual Method (MM), where regions of SAH blood were manually labeled in all slices of NCCT scans. To our knowledge, this comparative approach in 3D and 4D with clinical covariates has not been previously utilized in the literature. SAHVAI automation can help save precious time in this time-critical disease and stroke subtype, establishing a “ground truth” or “gold standard” for future SAH AI models as a benchmark for comparison. We acknowledge the limited size of this 3D/4D dataset and that it will require a larger validation set in the future.

The AI-driven SAHVAI model demonstrated superior speed, taking only seconds compared to over an hour for manual measurement of each SAH NCCT patient slice. This efficiency makes SAHVAI potentially useful for rapid evaluation of stroke patients, similar to established AI models for penumbral brain salvage in large vessel occlusion detection. SAHVAI measures SAHV blood quantitatively and quickly (6.7 seconds for 30 NCCT slices), comparable to existing commercial models like RAPID.AI, VIZ.AI, AIDoc, and Brainomix penumbra models. Our findings show that rapid SAHV correlates inversely with GCS, suggesting its potential as a biomarker of SAH severity.

Higher SAHV is inversely related to GCS and severity of illness, as indicated by the WFNS (World Federation of Neurological Surgeons) scale, and predicts longer LOS. Although this study is underpowered for predicting modified Rankin Scale outcomes, we believe SAHV can become a target for future interventions such as neuroprotective drugs or neurosurgical blood evacuation. The MM of SAHV measurement is very time-consuming, requiring over 60 minutes to manually segment each CT slice, which limits its practicality in routine practice. Consequently, our study′s cohort was limited to ten patients. In contrast, SAHVAI only took 6.7 seconds to measure the SAHV of 30 NCCT slices, with a calculated accuracy of 99.8%. In comparing our work to the literature review by Salman et al.^19^, we found no comparable AI model that combines SAH blood volume measurement speed, accuracy, and 3D (and 4D) mapping capabilities alongside clinical covariates. Other research teams have developed AI methods to detect SAH on NCCT scans and predict neurological events like DCI^24–29^. For instance, Nishi et al.′s AI model matched the accuracy of five neurosurgeons in detecting SAH and nonspecialists’ diagnostic accuracy^27^. In August 2023, Hu et al. proposed a U-Net deep-learning framework to identify and quantify SAH in NCCT scans, achieving a volume quantification in under 40 seconds with a precision of 72.2%. However, Hu et al.’s study did not establish a visualized 3D-brain mapping capability^25^.

Comparing our AI-enhanced approach and the Manual Method (MM), we observed similar downward SAHV curves over time, although they were not identical. In developing the MM, we added three extra-cisternal spaces (IPH, IVH, gyral/sulcal spaces) to the five cisternal spaces used in AI training. Including eight total spaces in the MM was justified by the belief that all brain blood matters and impacts SAH patients′ outcomes. However, the MM only measured the total SAHV of one CT scan, making it impossible to detect the additional three spaces quantitatively compared to the five cisternal spaces alone.

The differences between AI and manual SAHV measurement present a limitation, making direct quantitative comparison difficult. Most of the ten patients in our cohort had small to no IPH / IVH, so focusing on the five major cisternal spaces streamlined the model, enhancing its speed and accuracy for SAH blood. However, retraining the DL approach to include seven spaces requires a new software configuration, which our team will address in future projects.

Regarding the so-called 8^th^ space (gyral/sulcal space), manually labeling this tiny space proved impractical. A single cortical sulcal space with SAH blood often amounted to just a few pixels, typically totaling less than 1 cc. Training the AI SAHVAI model revealed it tended to overfit the 8^th^ sulcal space due to the brightness of SAH blood relative to the cortex. Given the minimal contribution of this space to the overall volumes in MM and the tradeoffs SAHVAI model performance, we excluded it, considering it a subject for future research.

Despite limitations, the AI and MM showed a mostly congruent decline in SAHV over time. SAHV might be more predictive of future clinical events like sVSP than the modified Fisher score (mFs), which was graded the same in all ten patients despite varied vasospasm severity. The SAHV-3D Brain Map could be more sensitive in vasospasm events and locations than mFs.

We found a day-by-day absolute SAHV difference of 5 ml between both methods, which seems to be sufficiently insignificant in clinical practice. Panchal et al. showed that small intracerebral hemorrhages (ICH) have lower mortality rates than massive ones, with 30ml being a cutoff for poorer outcomes^30^. The average relative % difference between both methods was 25.7% (0.26), which is not appreciable to the human eye per Brott et al^31^.

The difference in volume measurements could be due to SAH blood shifting into the spinal CSF space, which was not followed up on in our head CT scan focus. Patient records lacked details on posture, cardiac, and respiratory changes that might influence CSF distribution and SAHV. Further investigation is needed on the cerebrospinal fluid system, including the spinal cord^32^.

While data of our study were limited (n=10), linear regression suggested a trend towards increasing SAH blood volume with decreasing GCS score (R2=0.23, p=0.017) and increasing SAH blood volume with increasing LOS (R2=0.175, p=0.004). Sharma et al. found significant correlations between age, GCS score, and SAHV with discharge outcomes, DCI prediction, and in-hospital mortality.^16^

Further, Van der Steen et al. found that total blood volume (TBV) predicted poorer functional outcomes (determined with the mRS) and DCI in SAH more accurately than the mFS.^23,29^ Similar results were discovered by Yuan et al., where automated blood volume independently correlated with poorer clinical outcomes, whereas qualitative grading, such as mFS, does not. In addition to that, they discovered that cisternal and especially sulcal blood volume was most predictive for DCI and Vasospasm.^22^ Furthermore, Ramos et al. published their AI investigation and found total blood volume and volume location as factors of DCI occurrence alongside age, GCS, and treatment.^24^

More cases will be computed and analyzed to validate our approach at a greater database and, more precisely, to explore the correlation between volume and neurological outcomes at 30 days. Because the mRS was underpowered in the ten cases presented, it would be of high interest if a correlation could be found based on a greater database.

In conclusion, the introduction of SAHVAI-3D, a DL-driven NCCT-scan method, and the resultant “SAH 3D Brain map” may represent a new scientific tool to potentially transform the landscape for future SAH treatment and function as a baseline to compare patient outcomes. This innovation may fill a significant gap in current SAH imaging techniques and can open new avenues for translational researchers and clinicians to enhance patient care.

Although the SAHVAI-3D model needs to be explored and validated with a larger dataset, the “SAHVAI 3D Brain map”, given its preliminary findings, appears poised to become a new neurologic imaging biomarker in neurology, neurosurgery, and emergency medicine.

## Data Availability

All data produced in the present study are available upon reasonable request to the authors

## Disclosures

Drs Freeman and Erickson have a provisional patent on SAHVAI.

## Notes

### Competing Interest Statement

The authors have declared no competing interest.

### Funding Statement

This study did not receive any funding

### Author Declarations

Ethics committee/IRB of THE MAYO CLINIC waived ethical approval for this work

